# Risk of Alzheimer’s Disease is Associated with Longitudinal Changes in Plasma Biomarkers in the Multiethnic Washington Heights, Inwood Columbia Aging Project Cohort

**DOI:** 10.1101/2023.08.11.23293967

**Authors:** Yian Gu, Lawrence S. Honig, Min Suk Kang, Aanya Bahl, Danurys Sanchez, Dolly Reyes-Dumeyer, Jennifer J. Manly, Rafael A. Lantigua, Jeffrey L. Dage, Adam M. Brickman, Badri N Vardarajan, Richard Mayeux

**Affiliations:** Taub Institute for Research on Alzheimer’s Disease and the Aging Brain, Vagelos College of Physicians and Surgeons, Columbia University, New, York, New York, USA; G.H. Sergievsky Center, Vagelos College of Physicians and Surgeons, Columbia University, New York, New York, USA; Department of Neurology, Vagelos College of Physicians and Surgeons, Columbia University, and the New York Presbyterian Hospital, New York, New York, USA; Department of Epidemiology, Vagelos College of Physicians and Surgeons, Columbia University, and the New York Presbyterian Hospital, New York, New York, USA; Department of Medicine, Vagelos College of Physicians and Surgeons, Columbia University, and the New York Presbyterian Hospital, New York,New York, USA; Department of Pathology and Cell Biology, Vagelos College of Physicians and Surgeons, Columbia University, New York, New York, USA; Stark Neurosciences Research Institute, Indiana University School of Medicine, Indianapolis, Indiana, USA

**Author notes:** Correspondence: Richard Mayeux, MD, Neurological Institute of New York, 710 West 168^th^ Street, New York, NY 10032.

**Keywords:** amyloid, neurofilament light chain, tau, GFAP, blood biomarkers, Alzheimer’s disease, dementia, cognition, Hispanic

## Abstract

**INTRODUCTION:** Alzheimer’s disease (AD) biomarkers can help differentiate cognitively unimpaired (CU) individuals from mild cognitive impairment (MCI) and dementia. The role of AD biomarkers in predicting cognitive impairment and AD needs examination.

**METHODS:** In 628 CU individuals from a multi-ethnic cohort, Aβ42, Aβ40, phosphorylated tau-181 (P-tau181), glial fibrillary acid protein (GFAP), and neurofilament light chain (NfL) were measured in plasma.

**RESULTS:** Higher baseline levels of P-tau181/Aβ42 ratio were associated with increased risk of incident dementia. A biomarker pattern (with elevated Aβ42/Aβ40 but low P-tau181/Aβ42) was associated with decreased dementia risk. Compared to CU, participants who developed MCI or dementia had a rapid decrease in the biomarker pattern reflecting AD-specific pathological change.

**DISCUSSION:** Elevated levels of AD biomarker P-tau181/Aβ42, by itself or combined with a low Aβ42/Aβ40 level, predicts clinically diagnosed AD. Individuals with a rapid change in these biomarkers may need close monitoring for the potential downward trajectory of cognition.

**Research in Context:**

1. Systematic Review: Few studies have evaluated the clinical application of AD blood-based biomarkers longitudinally as antecedent risk predictors. Data from multiethnic populations are even more limited. How preclinical trajectories of blood-based biomarkers are related with the risk of developing clinically diagnosed MCI or AD is largely unknown.
2. Interpretation: High circulating level of P-tau181/Aβ42, by itself or combined with a low level of Aβ42/Aβ40, may predict development of incident clinical AD. Biomarkers levels of P-tau181, P-tau181/Aβ42, and NfL increase with age even among individuals who remain cognitively healthy. A rapid change in biomarkers may indicate the individuals in the active trajectory to develop clinically diagnosed MCI or AD.
3. Future Directions: Larger studies or meta-analyses are needed to examine whether the predictive utility of blood-based biomarkers for AD differs across racial/ethnic groups. Well-designed studies are needed to evaluate the optimal duration between repeated measures of biomarkers.

## 1. Background

Blood-based biomarkers for Alzheimer’s disease (AD), including β-amyloid (Aβ), tau, neurofilament light chain (NfL) and glial fibrillary acidic protein (GFAP), circulating molecular signatures of the amyloid, tau, and neurodegeneration (ATN) and inflammation, support their use in research and in clinical settings[1]. Compared to CSF and PET biomarkers [2, 3], blood-based biomarkers are less invasive, easily accessible, less expensive, and more suitable for large epidemiological studies and clinical trials. Potential clinical application of these blood-based biomarkers include application in the diagnosis, disease monitoring and prognosis, treatment management, screening, early detection, as well as risk prediction.

To date, much of the research on blood-based biomarkers has focused on their diagnostic value in research and in specialized settings [4–11]. Data from highly selected participants have been used for developing optimal thresholds for cut points, based on the presence or absence of AD-pathology in autopsy or amyloidosis in PET imaging as gold standards[12–15]. However, to date, no universially standardized and validated diagnostic cutpoints have been established, nor have blood-based biomarkers to been widely used to monitor the disease progression, or evaluate treatment responses. In addition to these applications, blood-based biomarkers could also aid in the identification of cognitively unimpaired (CU) individuals at risk of developing dementia. Blood-based biomarkers may also be of value as antecendent risk factors in prediction of MCI and AD and related dementia (ADRD) in asymptomatic individuals.

ADRD is known to have a long preclinical phase. Many of the neuropathological brain imaging changes occur during this preclinical stage. Thus, longitudinal changes in biomarkers levels in large, population-based, ethnically diverse cohorts would augment the value of blood-based biomarkers. Here, we examined whether blood-based biomarkers measured before the onset of clinical symptoms can predict the development of clinically diagnosed MCI or AD. This approach would help identify individuals at risk for disease-modifying treatments, augment studies examining biological mechanisms by identifying critical biomarker targets, and help identify modifiable factors that work through these biomarkers to delay the onset of the disease.

Using data from the Washington Heights, Inwood Columbia Aging Project (WHICAP) study, a longitudinal community-based, multiethnic population of older adults, we examined whether the initial measurement of blood-based biomarkers could predict subsequent MCI or AD diagnosis. We also investigated whether the rate of change in blood-based biomarkers over time differed among cognitively unimpaired individuals and those with newly diagnosed MCI or AD.

## 2. Methods

### 2.1. Study population

WHICAP is a multiethnic, community-based, prospective cohort study of clinical and genetic risk factors for dementia. Three waves of individuals were recruited in 1992, 1999, and 2009 in WHICAP, all using similar study procedures [16, 17]. Briefly, participants were recruited as representative of individuals living in the communities of northern Manhattan, 65 years and older, socioeconomically and racially diverse. At the study entry, each person underwent a structured interview of general health and function, followed by a comprehensive assessment including medical and neurological histories, standardized physical, neurological, and neuropsychological examinations. Individuals were followed every 18-24 months, repeating similar baseline examinations.

The institutional review boards of Columbia University gave ethical approval for this work. All participants provided written informed consent.

For this specific analysis, we selected individuals when they met the following criteria:1) indicated that they had not been diagnosed with AD or a related disorder at the initial interview; 2) had at least three blood samples at three different study follow-up visits 3) after each WHICAP follow-up visit had a clinical diagnosis of being cognitively healthy, MCI[18], or dementia [19]. For individuals whose diagnosis status changed over the WHICAP clinical follow-up visits, we selected plasma samples from the first visit, and any subsequent study visit. For participants remaining cognitively unimpaired through the follow-up, we chose blood samples collected at the first visit and all subsequent visits.

### 2.2. Cognitive assessment and clinical diagnosis of AD

At each WHICAP visit, individuals underwent a standardized neuropsychological battery [20] administered either in English or Spanish at baseline and each follow-up visit. Composite z-scores for four cognitive domains (memory, language/executive, speed, and visuospatial) were calculated based on a factor analysis using principal axis factoring and oblique rotation[20] on neuropsychological tests scores. The resulting factor structure and factor loadings were invariant across English and Spanish speakers[21].

All diagnoses were made in a diagnostic consensus conferences attended by a panel consisting of at least one neurologist and one neuropsychologist with expertise in dementia diagnosis, using results from the neuropsychological battery and evidence of impairment in social or occupational function. All-cause dementia which was determined based on *Diagnostic and Statistical Manual of Mental Disorders, 4th Edition criteria* [19]. Incident dementia was identified when the participants were clinically diagnosed with dementia for the first time during the follow-up study among those with a previous diagnosis with no dementia. For participants without dementia, MCI was assigned, as previously described[18], if the participant had memory complaint, had cognitive impairment in one or more cognitive domains, but with preserved activities of daily living. For all analyses, we combined MCI with dementia patients first and then examined the incident MCI and dementia separately when compared to CU.

### 2.3. Plasma biomarkers

Blood samples were collected by standard venipuncture in dipotassium ethylenediaminetetraacetic acid tubes. Plasma was prepared by centrifugation at 2,000xg for 15 minutes at 4°C within 2 hours after collection, aliquoted in polypropylene tubes, and frozen and stored at −80°C. Blood for DNA extraction was also collected, and apolipoprotein E (*APOE*) genotyping was performed at LGC Genomics and CD Genomics.

Plasma biomarker assays were performed between April 2022 and November 2022 using the single molecule array technology Quanterix Simoa (single molecule array)[22] HD-X platform (Quanterix, Billerica, MA, USA). Samples were diluted and assayed in duplicate per package insert instructions using three Quanterix kits: Neurology 3-Plex A (catalog No. 101995) for Aβ42, Aβ40, and T-tau; P-tau181 V2 Advantage (catalog No. 103714) for Tau phosphorylated at threonine 181 (P-tau181); and Neurology 2-Plex B (catalog No. 103520) for NfL and GFAP. Quantification functional lower limits for these analytes are 2.7 for Aβ40, 0.6 for Aβ42, 0.3 for T-tau, 0.3 for P-tau181, 0.8 for NfL, and 16.6 for GFAP, all in pg/mL. More than 5,000 assays were conducted for these analytes, and mean coefficients of variation are ≤ 5%. Ratios of Aβ42/Aβ40 and P-tau181/Aβ42 were calculated. Based on the literature, we *a priori* decided to focus on P-tau181 [23], neurodegeneration marker NfL [9, 24, 25], neuroinflammatory reactive astrogliosis marker GFAP[26], Aβ42/Aβ40 [27–29], and P-tau181/A β42 [30], while Aβ42, Aβ40, and t-tau were not investigated due to their limited value [31, 32].

### 2.4. Covariates

Demographic data including age (years), sex (male, female), ethnicity [white non-Hispanic, African American, Hispanic, and others], and education (years), were collected at the initial interview. *APOE*-ε4 genotype was defined based on the presence (either one or two) ε4 alleles. We calculated a modified Charlson Comorbidity Index including myocardial infarction, congestive heart failure, peripheral vascular disease, hypertension, chronic obstructive pulmonary disease, arthritis, gastrointestinal disease, mild liver disease, diabetes, chronic renal disease, and systemic malignancy, based on self-reported medical history and/or current medication use. Body mass index (BMI) was calculated as weight in kilograms divided by height in meters squared, with weight and height measured at the clinical visits, and was subsequently categorized into underweight or normal (<25 kg/m^2^), overweight (≥25 kg/m^2^ and <30 kg/m^2^), or obese (≥30 kg/m^2^).

In a subset sample of the study, we measured plasma creatinine using a kinetic colorimetric assay on an automated analyzer (Roche Integra 400 plus) at the Clinical Research Resource lab in the Irving Institute for Clinical and Translational Research, CUIMC. A creatinine level ≥1.3 mg/dl for men or ≥1.0 mg/dl for women was considered of an indication of renal dysfunction[33].

### 2.5. Statistical analysis

Descriptive statistics for individual demographic and clinical characteristics and plasma biomarker levels were compared among CU, incident MCI, and incident AD participants using χ^2^ for categorical variables and Kruskal-Wallis tests or ANOVA for continuous variables. Because the distributions of biomarkers were skewed, log-transformed biomarker levels were used in the analyses. For better visualize the biomarker levels, Z scores of the log-transformed biomarkers were used so the effect sizes could be compared among the biomarkers. Pearson’s correlations among the biomarkers as well as age, education, body mass index (BMI) were examined. Biomarker levels were also compared between men and women, *APOE*-ε4 carriers and non-carriers, and among race/ethnic groups using ANOVA.

We used COX proportional hazard models to examine whether baseline biomarker level could predict clinically diagnosed MCI and AD. Time variable was defined as the duration from the baseline to the last follow-up blood collection dates for controls, and the duration from the baseline to the incident MCI/AD diagnosis for those developed MCI/AD. Analyses were adjusted for age, education, sex, and ethnic group (model 1). In model 2, *APOE ɛ4* status, and Charlson Comorbidity Index were additionally adjusted. The individual biomarkers (pTau181, NfL, GFAP, Aβ42/Aβ40, and P-tau181/Aβ42) were included in COX models separately. Similar analyses were performed to examine the risk of incident MCI (incident AD was censored) and incident AD separately.

We used generalized estimating equations (GEE) models with repeated biomarker measures as outcomes to examine whether levels changed over time and whether individuals with incident MCI/AD and CU had different rates of change in plasma biomarkers. We used the duration from the baseline to the follow-up blood collection dates as the time variable. Models were adjusted with the same covariates as in the COX models. Similar analyses were performed to examine the difference between CU and incident MCI, and between CU and incident AD separately. Similar GEE models were also used to explore factors that are associated with rates of biomarker change over time among CU participants.

We performed supplementary analyses to assess the combined effects of biomarkers[5] as a predict of disease status. We performed principal component analysis (PCA) on the correlation matrix of NfL, GFAP, Aβ42/Aβ40, and P-tau181/Aβ42. The number of patterns to be retained was determined by eigenvalues >1.0, scree plot, parallel analysis, and interpretability of the factors. We performed the PCA at each visit separately. We considered biomarkers with an absolute factor loading value ≥ 0.30 on a pattern as dominant biomarker contributing to that biomarker pattern. The patterns derived from the three visit specific PCAs were similar, each having the first two patterns (PCA1 and PCA2) retained, which explained a total of 66%, 71%, and 71% variations of all the four biomarkers for visit 1, 2, and 3, respectively. For all visits, NfL (loadings 0.85), GFAP (loadings 0.81 to 0.87), and P-tau181/Aβ42 (loadings 0.36-0.48) had positive loadings for the first pattern (PCA1), while Aβ42/Aβ40 had a positive loading (loadings around 0.9) and P-tau181/Aβ42 had a negative loading (−0.3 to −0.7) for PCA2 (Supplementary Table 1). Each person received a pattern score (i.e., a linear combination of biomarker weighted by factor loadings) for each identified biomarker pattern. Thus, a higher PCA1 score would indicate a higher likelihood of neuronal injury, neuroinflammatory and neurodegenerative profile[34, 35], and a higher PCA2 score, in contrast, would indicate a lower likelihood of AD-specific pathological changes. We used the PCA1 and PCA2 scores in the above COX and GEE models to examine their predicting roles for AD and/or MCI.

Sensitivity analyses were performed to limit the GEE analysis to the pre-diagnosis visits only. Instead of using self-reported ethnic group information, we used genetic ancestry level in the analyses. We performed interaction analysis to examine whether the associations differed by ethnicity, sex, and *APOE ɛ4* status.

Two-sided statistical tests were conducted, and *p* < 0.01 with Bonferroni correction for multiple comparison (5 biomarkers: pTau181, NfL, GFAP, Aβ42/Aβ40, and P-tau181/Aβ42) was considered statistically significant. Statistical analyses were conducted with SPSS.

## 3. Results

### 3.1. Descriptive analysis

The current study included the first 628 cognitively unimpaired (CU) individuals selected from eligible WHICAP participants who met the above criteria. At visit two, which was on average 3.6 years from the baseline, 126 (20% of 628) converted from CU to MCI by clinical diagnosis and 16 (2.5%) converted to AD by clinical diagnosis and 486 (77%) remained to be CU; at visit three, which was 6.96 years from the baseline, additional 72 (15% of 486) CU had converted to MCI and 8 (1.6% of 486) converted to AD from CU, and 33 (26% of 126) MCI further converted to AD. Overall, 165 (26% of 628) individuals developed MCI, 57 (9%) developed AD, and 406 (65%) remained cognitively unimpaired during an average 6.96 (SD=3.07) years of follow-up. A total number of 585 (380, 151, 53 of CU, MCI, AD, respectively) had all three visits, but 43 (7%) (26, 13, 4 of CU, MCI, AD, respectively) had two samples only as one of their samples was degraded and could not be used to measure biomarker concentrations reliably.

The mean age of individuals at the initial visit was 73.4 (SD=5.6) years, 427 (67.9%) were women, and 20.4% carried one or two *APOE ɛ4* alleles. Individuals self-identified as NHW (27.7%), African American (25%), Hispanic (45.2%), or Others (2.1%).

Compared to CU individuals, those who developed either MCI or AD were older, were more likely to be Hispanic, had more comorbidities, and had higher levels of pTau181, NfL, GFAP, and a higher pTau181/Aβ42. There was no difference in the level of Aβ42/Aβ40 (Table 1).

**Table 1.** Characteristics of the study participants according to the disease outcome during follow-up.

|  | Cognitively<br>unimpaired<br>(N=406) | Incident MCI<br>(N=165) | Incident AD<br>(N=57) | Total<br>(N=628) | p |
| --- | --- | --- | --- | --- | --- |
| Age (years), mean (SD) | 71.8 (5.00) | 75.7 (5.04) | 78.2 (5.92) | 73.4 (5.58) | <0.0001 |
| Duration of storage time, mean (SD) | 15.54 (7.7) | 20.70 (9.58) | 21.97 (9.22) | 17.48 (8.75) | <0.0001 |
| Duration between visit 1 and 2, mean (SD) | 3.16 (1.64) | 4.40 (2.86) | 4.26 (2.29) | 3.58 (2.16) | <0.0001 |
| Duration between visit 1 and 3, mean (SD) | 6.35 (2.77) | 8.03 (3.41) | 8.21 (2.93) | 6.96 (3.07) | <0.0001 |
| Duration of follow-up time for disease outcome, mean (SD) | 6.16 (2.73) | 8.03 (4.45) | 6.91 (2.87) | 6.72 (3.38) | <0.0001 |
| Female, N (%) | 275 (67.7) | 112 (67.9) | 40 (70.2) | 427 (68.0) | 0.933 |
| Race/Ethnicity, N (%) |  |  |  |  | <0.0001 |
| Non-Hispanic White | 153 (37.7%) | 17 (10.3%) | 4 (7.0%) | 174 (27.7%) |  |
| Non-Hispanic Black | 112 (27.6%) | 39 (23.6%) | 6 (10.5%) | 157 (25.0%) |  |
| Hispanic | 129 (31.8%) | 108 (65.5%) | 47 (82.5%) | 284 (45.2%) |  |
| Others | 12 (3%) | 1 (0.6%) | 0 (0%) | 13 (2.1%) |  |
| APOE ε4 carrier, N (%) | 72 (17.7%) | 28 (17.0%) | 16 (28.1%) | 116 (18.5%) | 0.036 |
| Baseline biomarker levels: |  |  |  |  |  |
| P-tau181 (pg/mL), median [IQR] | 2.14 [1.65-2.90] | 2.16 [1.62-3.03] | 2.78 [1.86-3.71] | 2.22 [1.69-2.98] | 0.003 |
| Geometric mean (SD) | 2.187 (1.667) | 2.088 (2.090) | 2.770 (1.631) | 2.208 (1.790) | 0.005 |
| NfL (pg/mL), median [IQR] | 17.5 [13.1-24.7] | 19.8 [14.8-26.8] | 24.3 [16.7-30.6] | 18.2 [13.7-25.9] | <0.0001 |
| Geometric mean (SD) | 18.104 (1.625) | 20.203 (1.590) | 23.466 (1.620) | 19.077 (1.626) | <0.0001 |
| GFAP (pg/mL), median [IQR] | 138 [102-190] | 158 [112-215] | 193 [126-261] | 147 [106-206] | 0.001 |
| Geometric mean (SD) | 140.990 (1.666) | 155.200 (1.619) | 175.181 (1.693) | 147.470 (1.663) | 0.004 |
| AB42/AB40, median [IQR] | 0.044 [0.038-0.051] | 0.045 [0.041-0.052] | 0.044 [0.038-0.047] | 0.044 [0.039-0.0519] | 0.077 |
| Geometric mean (SD) | 0.044 (1.362) | 0.048 (1.577) | 0.044 (1.306) | 0.045 (1.421) | 0.014 |
| **P-tau181/Aβ42, median [IQR]** | 0.332 (0.536) | 0.365 (0.666) | 0.855 (3.15) | 0.387 (1.10) | <0.0001 |
| **Geometric mean (SD)** | 0.235 (1.966) | 0.244 (2.099) | 0.384 (2.312) | 0.248 (2.057) | <0.001 |
| **Serum Creatinine (mg/dl), mean (SD)** | 0.995 (0.347) | 1.21 (0.844) | 1.07 (0.341) | 1.05 (0.505) | 0.058 |
| **Charlson Comorbidity Index, mean (SD)** | 1.97 (1.45) | 2.34 (1.50) | 2.75 (1.53) | 2.13 (1.49) | <0.0001 |
| **BMI, N (%)** |  |  |  |  | 0.16 |
| **Underweight or normal** | 81 (20.0%) | 22 (13.3%) | 7 (12.3%) | 110 (17.5%) |  |
| **Overweight** | 141 (34.7%) | 51 (30.9%) | 20 (35.1%) | 212 (33.8%) |  |
| **Obese** | 73 (18.0%) | 40 (24.2%) | 12 (21.1%) | 125 (19.9%) |  |

At baseline, there were strong positive correlations among P-tau181, NfL, GFAP, and P-tau181/Aβ42, but they were not correlated with Aβ42/Aβ40 except for the negative correlation between P-tau181 and Aβ42/Aβ40 (Table 2). At baseline, those who were older and those who had more comorbidities had higher biomarkers levels, with the exception of Aβ42/Aβ40 (Table 2). Women had higher levels of NfL and GFAP than men. *APOE ɛ4* carriers had higher levels of pTau181 and a higher P-tau181/Aβ42 than non-carriers. Hispanics had lower NfL and pTau181 than white non-Hispanics or African American individuals.

**Table 2.** Correlations among biomarkers and demographic factors.

| | Ptau181 | NfL | GFAP | Aβ42/Aβ40 | Ptau181/Aβ42 |
| --- | --- | --- | --- | --- | --- |
| P-tau181 | 1.0 |  |  |  |  |
| NfL | .350** | 1.0 |  |  |  |
| GFAP | .218** | .502** | 1.0 |  |  |
| Aβ42/Aβ40 | -.189** | -0.001 | -0.002 | 1.0 | |
| p-tau181/Aβ42 | .597** | .249** | .130** | -0.044 | 1.0 |
| Age, years | .189** | .388** | .332** | 0.008 | .201** |
| Charlson Comorbidity index | .182**,a | .211**,a | .092* | -.018 | .124**,a |
| BMI, kg/m<sup>2</sup> | .009 | -.046 | -.022 | -.076 | .061 |

Log-transformed values of biomarkers were used in the Pearson correlation analyses. \*p<0.05;
\*\* p<0.01; a p<0.01 after adjusted for age.

When examining the repeated measures of biomarkers among CU participants, levels of P-tau181 (b=0.009, p<0.001), NfL (b=0.016, p<0.001), and GFAP (b=0.013, p<0.001) increased during follow up, adjusting for baseline age, sex, ethnicity, and education (Figure 1). Women had slower increase in P-tau181 (β for interaction female*time=-0.012, p=0.002), and African American and Hispanic individuals had faster increase than white non-Hispanics in NfL (β for Hispanic*time=0.010, p=0.009; β for African American*time=0.014, p=0.007), and PAC1 (β for Hispanic*time=0.041, p=0.007; β for African American*time=0.04, p=0.034).

**Figure 1.**
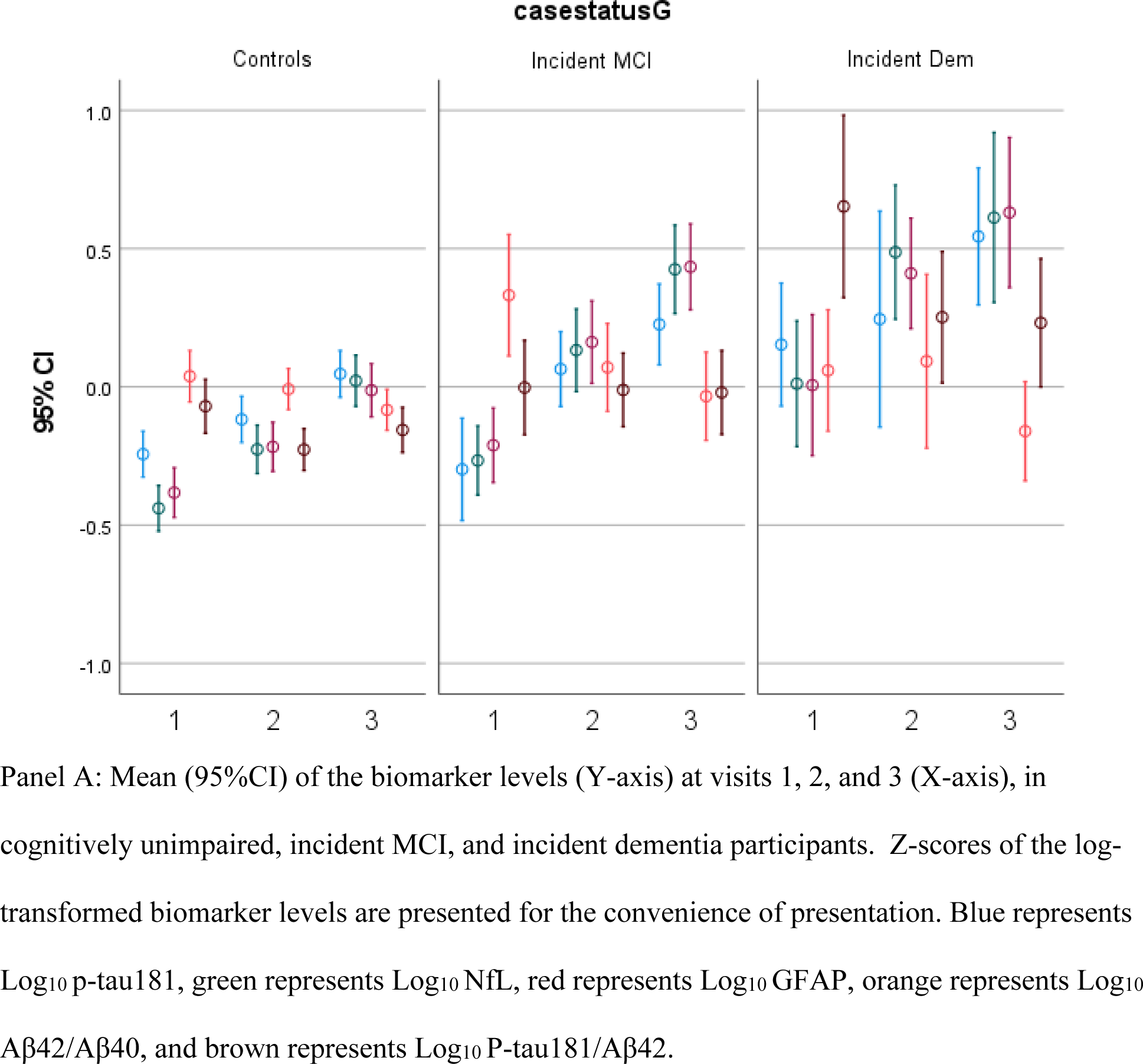
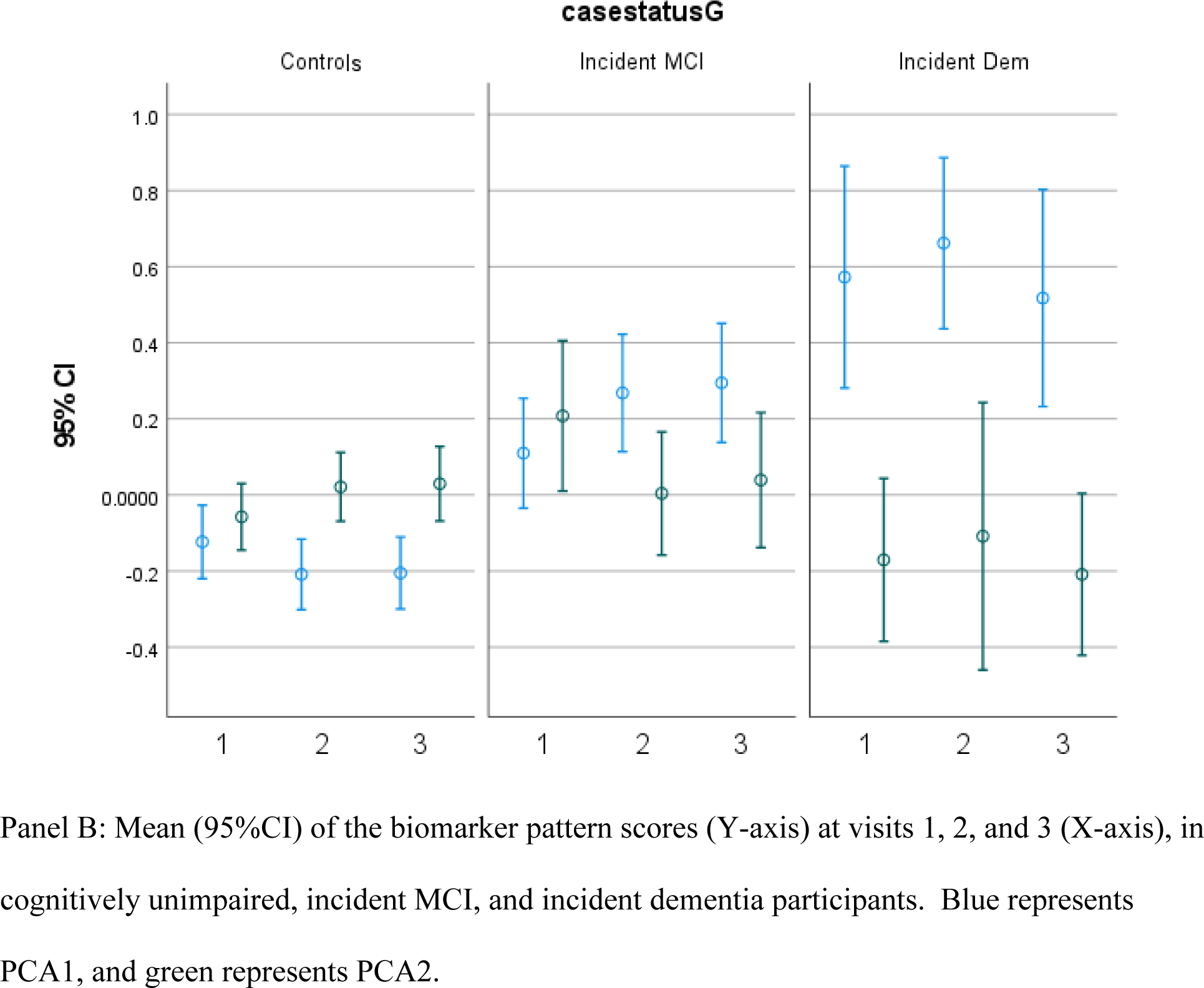
Blood-based biomarkers of Alzheimer’s disease at different visits, among cognitively unimpaired, incident MCI, and incident AD.

### 3.2. Longitudinal association of the baseline blood-based biomarkers with clinically diagnosed incident MCI/AD

In COX models adjusted for age, sex, ethnic group, and education (Table 3, Model 1), we found higher baseline level of P-tau181 (HR=4.77, 95%CI=1.52-14.95, p=0.007) and Ptau181/Aβ42 ratio (HR=2.94, 95%CI=1.50-5.78, p=0.002) were associated with increased risk of developing incident AD by the clinical diagnosis. Additionally adjusting for *APOE ɛ4* status and Charlson comorbidity index, the significant association remained for P-tau181/Aβ42 (HR=3.13, 95%CI=1.43-6.87, p=0.004) but the association for P-tau181 was attenuated (HR=2.88, 95%CI=0.79-10.56, p=0.11) (Table 2, Model 2). While other biomarkers did not reach significance, their associations with AD risk were all in the expected direction (Table 3, Model 2). In the supplementary analyses, the PCA1 (HR=1.50, 95%CI=1.12-2.01, p=0.006, Table 2, model 1) and PCA2 (HR=0.66, 95%CI=0.49-0.88, p=0.005, Table 2, model 1) were both associated with incident AD in Model 1, and similar results were found in Model 2.

**Table 3.** Cox proportional hazards models.

|  | Model 1 |  | Model 2 |  |
| --- | --- | --- | --- | --- |
| *Inc. MCI+AD* | HR (95%CI) | *P* | HR (95%CI) | *P* |
| P-tau181 | 1.06 (0.93-1.21) | 0.37 | 1.51 (0.88-2.59) | 0.135 |
| NfL | 1.12 (0.94-1.33) | 0.194 | 1.29 (0.57-2.92) | 0.548 |
| GFAP | 1.10 (0.95-1.28) | 0.211 | 1.72 (0.82-3.61) | 0.151 |
| Aβ42/Aβ40 | 0.92 (0.81-1.05) | 0.206 | **0.24 (0.09-0.62)** | **0.003** |
| P-tau181/Aβ42 | 0.93 (0.81-1.06) | 0.289 | 0.75 (0.47-1.19) | 0.217 |
| PCA1 | 1.06 (0.92-1.23) | 0.426 | 1.04 (0.88-1.23) | 0.630 |
| PCA2 | 0.86 (0.76-0.99) | 0.029 | 0.84 (0.73-0.97) | 0.021 |
| *Inc. MCI* | HR (95%CI) | *P* | HR (95%CI) | *P* |
| P-tau181 | 1.06 (0.93-1.21) | 0.37 | 1.22 (0.67-2.23) | 0.508 |
| NfL | 1.12 (0.94-1.33) | 0.194 | 1.73 (0.73-4.13) | 0.215 |
| GFAP | 1.10 (0.95-1.28) | 0.211 | 1.69 (0.79-3.63) | 0.178 |
| Aβ42/Aβ40 | 0.92 (0.81-1.05) | 0.206 | 0.67 (0.24-1.85) | 0.440 |
| P-tau181/Aβ42 | 0.93 (0.81-1.06) | 0.289 | 0.67 (0.39-1.16) | 0.156 |
| PCA1 | 1.07 (0.91-1.27) | 0.417 | 1.08 (0.90-1.30) | 0.401 |
| PCA2 | 0.95 (0.82-1.09) | 0.453 | 0.99 (0.85-1.15) | 0.885 |
| *Inc. AD* | HR (95%CI) | *P* | HR (95%CI) | *P* |
| P-tau181 | **4.77 (1.52-14.95)** | **0.007** | 2.88 (0.79-10.56) | 0.110 |
| NfL | 3.42 (0.81-14.48) | 0.095 | 3.04 (0.58-15.89) | 0.189 |
| GFAP | 3.49 (0.92-13.26) | 0.067 | 3.44 (0.79-14.96) | 0.099 |
| Aβ42/Aβ40 | 0.10 (0.01-0.81) | 0.031 | 0.07 (0.01-0.79) | 0.031 |
| P-tau181/Aβ42 | **2.94 (1.50-5.78)** | **0.002** | **3.13 (1.43-6.87)** | **0.004** |
| PCA1 | **1.50 (1.12-2.01)** | **0.006** | 1.49 (1.07-2.07) | 0.019 |
| PCA2 | **0.66 (0.49-0.88)** | **0.005** | **0.63 (0.45-0.88)** | **0.006** |

- PCA1 and PCA2 were derived from principal component analysis using log-transformed values of A β42/A β40, P-tau181/AB42, NfL, and GFAP, with PCA1 having positive loadings on P-tau181/AB42, NfL, GFAP, and PCA2 having positive loading on A β42/A β40 and negative loading on pTau181/AB42.

### 3.3. Longitudinal analyses to examine whether the rate of blood-based biomarkers change over time differs in cognitively healthy older adults and incident MCI/AD patients

We found a relatively faster increase of P-tau181, NfL, GFAP, and P-tau181/Aβ42 and faster decrease of Aβ42/Aβ40 in incident MCI/AD compared to CU participants; however, the results were not significant (Table 4). Nevertheless, incident MCI/AD participants had a different rate of change in PCA2 compared to CU participants [β=-0.034 (−0.060 - −0.008), p=0.010], adjusted for age, sex, ethnic group, and education, *APOE* status, and comorbidity score (Table 3, Model 2). Furthermore, similar results were found comparing incident MCI [β= −0.036 (−0.064 - −0.009), p=0.010] to CU.

**Table 4.** Longitudinal change of biomarkers in relation to incident MCI and AD.

|  | Model 1 |  |  |  | Model 2 |  |  |  |
| --- | --- | --- | --- | --- | --- | --- | --- | --- |
|  | 95%CI |  |  |  | 95%CI |  |  |  |
| *Inc. MCI or AD vs CU* | B | Lower | Upper | P | B | Lower | Upper | P |
| P-tau181 | 0.004 | -0.002 | 0.010 | 0.202 | 0.005 | -0.002 | 0.011 | 0.156 |
| NfL | 0.002 | -0.003 | 0.007 | 0.368 | 0.002 | -0.004 | 0.007 | 0.575 |
| GFAP | 0.003 | -0.002 | 0.008 | 0.258 | 0.003 | -0.003 | 0.008 | 0.335 |
| Aβ42/Aβ40 | -0.003 | -0.006 | 0.000 | 0.080 | -0.004 | -0.007 | 0.000 | 0.055 |
| P-tau181/AB42 | 0.003 | -0.003 | 0.010 | 0.343 | 0.003 | -0.004 | 0.010 | 0.358 |
| PCA1 | 0.022 | 0.001 | 0.043 | 0.040 | 0.016 | -0.007 | 0.039 | 0.181 |
| PCA2 | **-0.032** | **-0.055** | **-0.009** | **0.007** | **-0.036** | **-0.062** | **-0.011** | **0.005** |
|  | 95%CI |  |  |  | 95%CI |  |  |  |
| *Inc. dem vs CU* | B | Lower | Upper | P | B | Lower | Upper | P |
| P-tau181 | 0.0004 | -0.008 | 0.009 | 0.919 | -0.001 | -0.009 | 0.007 | 0.832 |
| NfL | 0.003 | -0.007 | 0.012 | 0.569 | 0.001 | -0.010 | 0.011 | 0.903 |
| GFAP | 0.006 | -0.002 | 0.014 | 0.151 | 0.005 | -0.004 | 0.014 | 0.266 |
| Aβ42/Aβ40 | -0.003 | -0.007 | 0.002 | 0.263 | -0.002 | -0.008 | 0.003 | 0.358 |
| P-tau181/AB42 | -0.005 | -0.016 | 0.006 | 0.372 | -0.005 | -0.018 | 0.008 | 0.416 |
| PCA1 | 0.013 | -0.024 | 0.050 | 0.498 | 0.002 | -0.040 | 0.045 | 0.910 |
| PCA2 | -0.028 | -0.063 | 0.007 | 0.119 | -0.027 | -0.068 | 0.014 | 0.200 |
|  | 95%CI |  |  |  | 95%CI |  |  |  |
| *Inc. MCI vs. CU* | B | Lower | Upper | P | B | Lower | Upper | P |
| P-tau181 | 0.005 | -0.002 | 0.012 | 0.159 | 0.006 | -0.001 | 0.014 | 0.087 |
| NfL | 0.002 | -0.003 | 0.008 | 0.439 | 0.002 | -0.004 | 0.008 | 0.537 |
| GFAP | 0.002 | -0.004 | 0.008 | 0.479 | 0.002 | -0.004 | 0.008 | 0.508 |
| Aβ42/Aβ40 | -0.003 | -0.006 | 0.001 | 0.109 | -0.004 | -0.008 | 0.000 | 0.055 |
| P-tau181/AB42 | 0.006 | -0.001 | 0.012 | 0.114 | 0.006 | -0.001 | 0.013 | 0.105 |
| PCA1 | 0.025 | 0.002 | 0.047 | 0.033 | 0.020 | -0.005 | 0.044 | 0.115 |
| PCA2 | **-0.033** | **-0.058** | **-0.008** | **0.010** | **-0.039** | **-0.067** | **-0.012** | **0.005** |

- B values in the table indicate the beta coefficient for the interaction between the disease status\*time, with time being the duration (years) between the first blood visit to the follow-up blood visits. Significant interactions indicate the rate of biomarker change over time in incident MCI and/or AD patients differ from the rate in cognitive normal participants.

### 3.4. Sensitivity analysis

The GEE analyses results did not change when limiting analyses to the pre-diagnosis visits only, i.e. excluding the third visits of 94 individuals who had already developed MCI or dementia at the second visit, with a rapid decrease in PCA2 comparing incident MCI/AD [β= −0.034 (95%CI: −0.06, −0.008), p=0.010] to CU adjusted for age, sex, ethnicity, education, APOE status, and comorbidities (Model 2).

Adjusting for the ancestry level (three principal component factors representing ancestry level of whites, blacks, and Hispanics) instead of self-reported race/ethnicity, the results were similar. COX still showed baseline P-tau181/Aβ42 ratio [HR=3.26 (95%CI=1.43-7.41, p=0.005)] and PCA2 pattern [HR=0.64 (95%CI=0.46-0.88, p=0.008)] and both significantly predicted incident dementia. GEE also showed incident MCI had faster decline [b=-0.038, 95%CI=(−0.065, −0.010), p=0.008] in PCA2 biomarker pattern compared to CU after adjusting for age, sex, ancestry level, education, *APOE* status, and comorbidities.

We found sex, ethnicity, or *APOE ɛ4* did not modify the association of biomarkers and disease outcome (p>0.10 for all interaction terms) (data not shown).

In the subset of the study population (N=251), we found incident MCI/AD had a faster decline [b=-0.048, 95%CI= (−0.080, −0.016), p=0.003] in PCA2 compared to CU after adjusting for Model 2 covariates (age, sex, ethnicity, education, *APOE* status, and comorbidities) as well as creatinine and BMI.

## Discussion

In this community-based cohort of cognitively unimpaired adults, we found higher level of P-tau181/Aβ42, and a biomarker pattern of higher level of P-tau181/Aβ42 along with lower level of Aβ42/Aβ40 (i.e. PCA2), predicted the development of incident clinical AD. In addition, those who developed MCI/AD had a rapid decrease in Aβ42/Aβ40 along with increase P-tau181/Aβ42, compared to participants remained cognitively unimpaired.

### Predictive value of single measure of biomarkers

Our results provide important evidence that blood-based AD biomarkers have clinical utility in predicting incident MCI and AD and in monitoring the cognitive trajectory among cognitively unimpaired participants. We found among all biomarkers, P-tau181 or P-tau181/Aβ42 were the biomarkers most strongly associated with risk of cognitive impairment, consistent with previous studies that found baseline P-tau181 predicting AD risk or cognitive decline in cognitively unimpaired individuals[4, 24, 36]. Although generally studies found biomarkers of the A/T/N and X (inflammation, etc.) framework are associated with increased risk of dementia, results for individual biomarkers other than P-tau181 are not always consistent. In 300 participants of an Amsterdam study [37], both GFAP and Aβ42/Aβ40, but not NfL, were independently associated with incident dementia. In another study, GFAP showed the best performance, followed by NfL and P-tau181, in predicting clinical AD risk[38]. In the Rotterdam study[39], baseline NfL, Aβ42, and Aβ42/Aβ40 ratios, but not Aβ40 or T-tau, were associated with risk of developing dementia. Overall, there is no consensus with regard to the relative importance of the biomarkers in predicting AD risk, but all the significant findings reported by various studies are in the expected direction, i.e. increased biomarkers (or decrease Aβ42/Aβ40) are associated with increased risk of AD. Differences in sample size, age, sex, race/ethnicity, and comorbidities, factors that may influence the biomarker levels, as found in the current and other studies[13], may partially explain the inconsistent findings across studies. Recent studies have evaluated the dynamic changes of the biomarkers along the AD continuum, and found GFAP may be an early AD biomarker, while p-tau181 and NfL may subsequently predict AD at a later time [40, 41]. Thus, inconsistent results from different studies might also be due to the different timing of blood sample collection for biomarker measurements.

### Repeated measure of biomarkers

While biomarker levels in a one-time measurement may help identify individuals at high risk of developing AD, monitoring the trajectory of biomarkers by repeated measurements might provide additional predictive value at an even earlier stage. An increase in the biomarker levels may indicate the beginning of the pathological process, and thus may provide a critical window for effective early prevention[42]. We found all biomarkers, except for Aβ42/Aβ40, increased over time within individuals, consistent with the cross-sectional findings of positive correlation between age and these biomarkers in the current study, as well as findings in previous studies that reported similar increase of biomarkers over time [25, 36, 37, 39, 43–45]. However, we did not find a significant difference in the rate of change of the biomarkers comparing CU and those who developed cognitive impairments during follow-up. Data are scarce in examining the rate of change of the biomarkers in relation to clinical disease status. In the Mayo Clinic Study of Aging (MCSA) study, the rate of increase in plasma NfL was not different between CU and MCI [25]. In contrast, studies found mean plasma NfL levels, but not Aβ42[39] or GFAP[37], increased faster in participants who developed dementia compared to participants who remained dementia-free[45, 46]. Additional evidence also supports the increase in plasma NfL over time may indicate an active trajectory to MCI or AD. For example, an increase in NfL was associated with increasing level of amyloid PET [25] and faster cognitive decline[25, 45]. Although we did not find rate of NfL change varied between CU and MCI/AD, NfL did have a large contribution to the biomarker pattern PCA1, which increased at a marginally significantly faster speed in incident MCI than in CU.

Monitoring the change of biomarkers other than NfL might also be important. Longitudinal changes of plasma P-tau181 was found to be steeper in MCI than in CU[36], and was also associated with cognitive decline[24]. In addition, increase in plasma P-tau181 was related to the decrease in gray matter volume in certain brain areas [4, 47] or amyloid deposition in the brain[48], which might stand as mediators leading to cognitive decline and dementia[24]. In the current study, the biomarker pattern PCA2, with P-tau181 and Aβ42/Aβ40 as the key components, showed different changes in MCI/AD compared to CU.

Overall, the biomarkers tended to have more rapid change among those developing MCI, but not those developing AD, compared to CU. One possible reason could be the biomarkers were already high at baseline, thus may be closer to the ‘ceiling’ and therefore slower change, in the AD patients[41].

### Combination of biomarkers

We found that most of the biomarkers were associated with the outcome in the expected direction, although after correction for multiple testing, some not statistically significant. Studies found some AD biomarkers can provide non-overlapping information on neuropathological changes[49], suggesting a holistic evaluation of the combined effect of the biomarkers may better capture the overall ATN and inflammation profile of an individual. Indeed, we found two patterns performed better than individual biomarkers in predicting incident dementia, and repeated measures of the patterns, but not the individual biomarkers, could help monitor development of MCI. Few previous studies combined multiple biomarkers[39, 50, 51]. Similar to the pattern PCA2 in our study, the Rotterdam study[39] found combining the lowest quartile group of Aβ42 with the highest of NfL resulted in a stronger association with dementia, compared to the highest quartile group of Aβ42 and lowest of NfL. In another study, researchers found that combining Aβ42/Aβ40 and plasma GFAP, with age and *APOE* status, provided the optimal panel identifying a positive amyloid status[50]. A recent study also reported a stronger association with incident dementia for joint NfL and GAFP compared to either of the two individual biomarkers[51]. Overall, while there is no consensus of the best combination of the biomarkers in predicting cognitive impairment or making diagnosis, these studies point to an increased value of examining the biomarkers simultaneously, as compared to individual ones, in dementia research. As AD is known to be a complex multi-factorial neurodegenerative disorder[52], combining biomarkers measuring different pathways may indeed be necessary in future studies.

### Limitations and advantages

Most studies use amyloid PET or autopsy to develop optimal threshold cut points for diagnosis when using biomarkers. However, here we used the full range of each biomarker to assess risk of developing clinically diagnosed MCI or AD. Here we show that there is a linear relationship beween increased P-tau181 or P-tau181/Aβ42 ratio, which would not be confirmed if a dictomous cut point had been used. Thus, there is a clear disadvantage in using derived cut points to assess risk of disease because there is a loss of potentially valuable information. As a risk biomarker using analyWe did not measure other P-tau isoforms (p-tau217, p-tau231, p-tau205, p-tau212) because they were not commerically available when the study began. They may have shown significant associations with cognitive decline in non-demented subjects [53, 54]. However, P-tau isoforms have moderate-to-strong correlation to each other. While we adjusted for multiple key factors including age, sex, ethnicity, and APOE, we did not have other putative confounders, such as creatinine and BMI, in the entire study population. However, to be consistent with the literature[55], we adjusted for these variables in the subset but with similar results. Although this study is relatively large and has repeated measures of biomarkers, only a small number of white non-Hispanic and African American individuals developed AD, thus our statistical power to detect significant results in those ethnic groups was limited.

Our study has many strengths. Our study population was from a community-based, multiethnic cohort, thus may have good representation of general population. We measured both biomarkers and outcome longitudinally, with three measures of biomarkers in most participants and the follow up time up to 23 years. We adjusted for potential confounders. In addition to examining individual biomarkers, we derived two biomarker patterns, which were quite robust across different visits and showed stronger association with outcomes than individual biomarkers.

While many studies use autopsy or PET imaging to establish optimal thresholds or cut points for the diagnosis of AD, there is still no universal or established cut points for the use of these AD biomarkers as diagnostics. However, in this investigation, we found that AD biomarkers collected longitudinally may be clinically useful as adjuncts to the neurological and cognitive evaluations. Previous cross-sectional studies have concluded that these AD biomarkers provide a physiological basis for the diagnosis of AD consistent with the A/T/N recommendations. Here we did not determine, nor did we include thresholds or cut points, rather we used the AD biomarkers to determine whether they are consistent with the clinical diagnosis. Advances in therapeutic strategies for AD need to include risk prediction. The AD biomarkers used here represent a reasonable approach to risk prediction.

## Data Availability

All data produced in the present study are available upon reasonable request to the authors.

## Acknowledgments

Data collection and sharing for this project was supported by the Washington Heights-Inwood Community Aging Project (WHICAP; PO1AG007232, R01AG037212, RF1AG054023, AG037212, AG072474, AG066107, AG059013) funded by the National Institute on Aging (NIA). This manuscript has been reviewed by WHICAP investigators for scientific content and consistency of data interpretation with previous WHICAP Study publications. We acknowledge the WHICAP study participants and the WHICAP research and support staff for their contributions to this study.

## Notes

### Competing Interest Statement

The authors have declared no competing interest.

### Funding Statement

This study was funded by the National Institute on Aging grants PO1AG007232, R01AG037212, RF1AG054023, AG037212, AG072474, AG066107, AG059013, and by the National Center for Advancing Translational Sciences, National Institutes of Health, through grant No. UL1TR001873.

### Author Declarations

The institutional review boards of Columbia University gave ethical approval for this work.

