## Supplemental Table 1 for "Risk of Alzheimer’s Disease is Associated with Longitudinal Changes in Plasma Biomarkers in the Multiethnic Washington Heights, Inwood Columbia Aging Project Cohort"

Supplementary Table 1. Biomarker patterns derived from principal component analyses.

| | visit 1 | | visit 2 | | visit 3 | |
|---|---|---|---|---|---|---|
| | PCA1 | PCA2 | PCA1 | PCA2 | PCA1 | PCA2 |
| Aβ42/40 ratio | | 0.957 | | 0.872 | | 0.868 |
| P-tau181/Aβ42 ratio | 0.482 | -0.303 | 0.367 | -0.647 | 0.356 | -0.683 |
| NfL | 0.854 | | 0.853 | | 0.835 | |
| GFAP | 0.808 | | 0.863 | | 0.867 | |

The patterns were derived from three visit-specific principal component analyses, each having the first two patterns (PCA1 and PCA2) retained. Loadings less than 0.3 were omitted in the table.
